# Left Ventricular Volume and Function Assessment Using a Reduced-Slice Approach in Cardiovascular Magnetic Resonance

**DOI:** 10.64898/2026.05.29.26354413

**Authors:** Aditya Tejaswi, Alexander Fyrdahl, Andreas Sigfridsson

**Affiliations:** Department of Molecular Medicine and Surgery, Karolinska Institutet, Stockholm, Sweden; Unit of Clinical Physiology, Karolinska University Hospital, Stockholm, Sweden

**Keywords:** Left Ventricle, Volumetry, Stroke Volume, Ejection Fraction, Scan Time Reduction, Global Function

## Abstract

**Object:** To test the hypothesis that a reduced-slice geometric model can maintain left ventricular (LV) volumetry, thereby providing a rapid and reliable clinical alternative to conventional full-stack planimetry.

**Materials and Methods:** In a retrospective cohort of 342 patients with full-stack cardiovascular magnetic resonance (CMR), nine geometric combinations (3, 4, or 5 short-axis (SAx) slices and a 2, 3, or 4 chambered long-axis (LAx) slice) were evaluated by decimating the original acquisitions. Volumes were calculated using trapezoidal mid-cavity approximations and a generalized prismoidal model for the basal region. This was compared to the full-stack reference using concordance correlation coefficient (CCC), repeated measures ANOVA, pairwise tests and Bayes factor log_10_(BF_10_) analysis.

**Results:** The choice of LAx view influenced the accuracy approximately 50 times more than SAx slice count. For a 2ch + 5 SAx protocol, EF bias was just 0.83% (LoA: -6.18 to 7.84%), with a minimum detectable change (MDC) of 7.01%, compared to 8.7% reported for expert human readers, suggesting strong concordance (CCC>0.90) with full-stack. Bayesian analysis decisively supported 5 over 3 SAx slices (log_10_(BF_10_) = 6.42).

**Conclusion:** This proposed model successfully minimizes scan time and breath-holds while maintaining clinical accuracy. Functional metric bias remains lower than scan-rescan variability.

## 1 Introduction

Cardiovascular Magnetic Resonance (CMR) is the current gold standard for quantifying left-ventricular (LV) functions through volumetric measurements such as End Diastolic Volume (EDV), End Systolic Volume (ESV), Stroke Volume (SV) and Ejection Fraction (EF). Existing CMR methods involve acquisition of a cine short axis (SAx) stack of slices, along with the 2, 3 and 4-chamber long axis (LAx) views (1) acquired across multiple breath holds. The SAx stack typically covers the entire left ventricle of the heart with contiguous slices from base to apex and can include anywhere from 9 to 15 slices depending on the size of the heart. The scan time, and the number of breath holds, is therefore directly linked to the number of acquired slices. Analysis time also increases with the number of slices, as the reader has to contour the endocardial and epicardial borders in every slice to obtain the EDV and ESV, from which the EF and SV are calculated. Manual contouring is notably prone to inter-and-intra observer variability (2), but also variability at the apical and basal slices due to the longitudinal shortening of the heart in systole. The uncertainty in the selection of the basal slice has been shown to cause up to a 13% discrepancy in volumetric measurements between different observers (3). While artificial intelligence (AI) has accelerated image analysis, the duration required for full-stack image segmentation remains a primary rate-limiting step in the clinical workflow.

Longitudinal follow-up may be confounded by the variability in manual contouring (4), (5), and challenges arise in assessing whether the difference is due to disease progression or due to the inter or intra-observer variability (6), (7), (8).

One approach to reduce the acquisition time for CMR volume estimation is to acquire and approximate the ventricular volume from fewer slices. A combination of a long axis image with few short axis images has shown to approximate the full stack volume remarkably well (9) despite not accounting for the basal most volume between the most basal short axis slice and the mitral annulus. The current study improves upon previous work by adding the sub-annular volume to the approximation. The aim of this work is to present and evaluate the proposed method in a clinical cohort and to determine the accuracy and tradeoffs from different selections of short axis and long axis slices, respectively. We hypothesize that the reduced-slice method provides acceptable accuracy compared to full stack reference, but with a reduced number of breath holds.

## 2 Materials and Methods

### 2.1 Subjects

To evaluate the proposed reduced-slice volume calculation method, we retrospectively identified 582 adult patients, without arrhythmia, undergoing CMR examinations at the Karolinska University Hospital between September 5 2024 and June 25 2025. Data from these patients were retrieved from the PACS. Data collection from these patients was approved by the Swedish Ethical Review Authority (DNR: 2011/1077-31/3 and 2022-01151-02), and all patients provided written informed consent. An additional 81 patients were excluded for image quality issues or incomplete exams, and 159 patients who didn’t meet the minimum criteria of five intracavitary slices at end-systole (see Fig. 1). The final study cohort comprised 342 patients, 54 (40 – 65) years, 48.4% females, as shown in Table 1. All examinations were performed at 1.5T (MAGNETOM Sola, Siemens Healthcare, Forchheim, Germany), and included a cine short-axis scan, a two-chamber, three-chamber and a four-chamber long-axis scans acquired under breath-hold. Automatic left-ventricular segmentation was performed using the AI Automate feature of the freely available software Segment 4.1.0.1 (Medviso, Lund, Sweden) (10).

**Figure 1:**
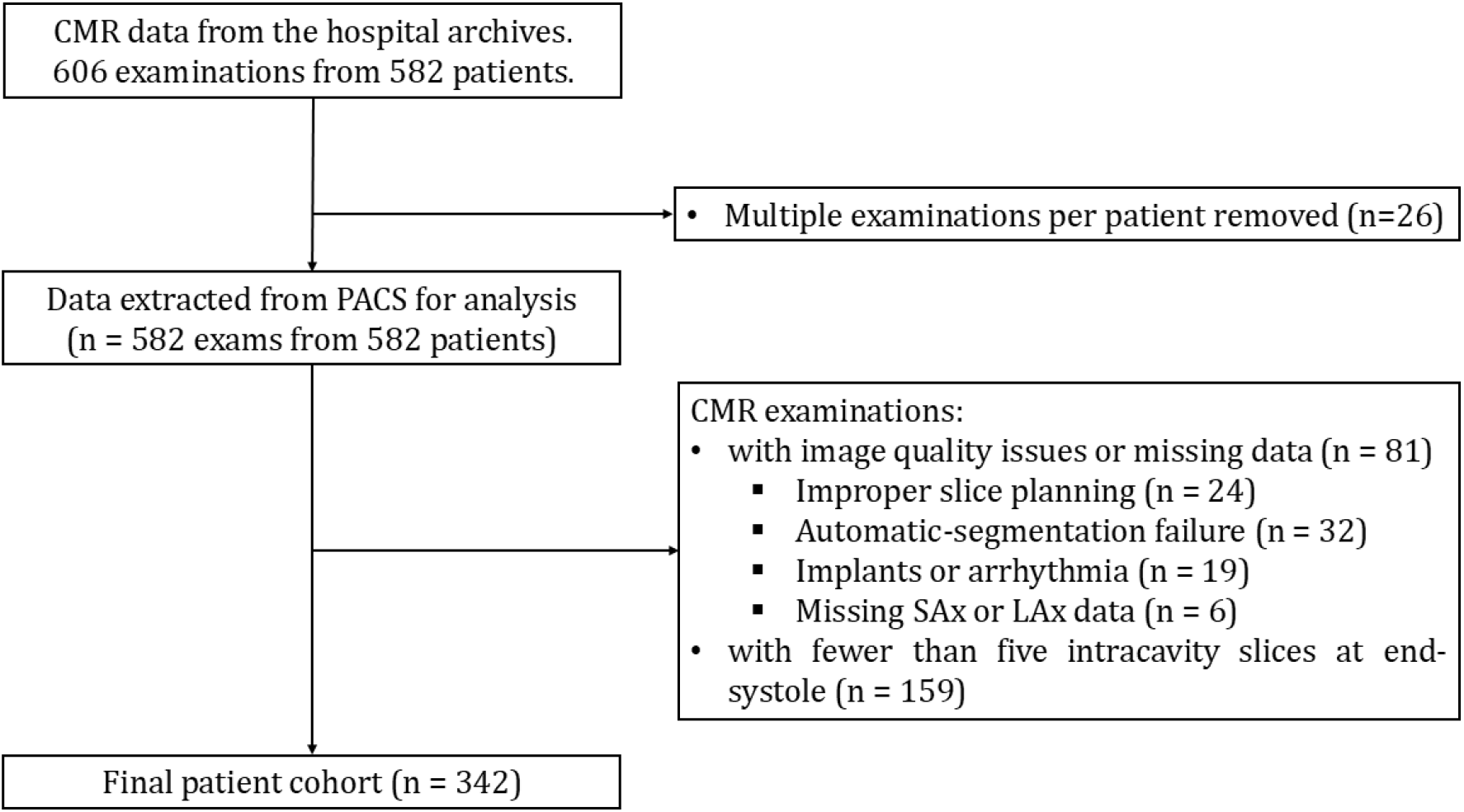
Selection of study cohort. The figure displays the inclusion and exclusion of CMR patient data for this study. *Intracavity slices refer to the slices between the mitral annulus and apex across all cardiac phases.

**Table 1:** Patient characteristics and imaging metadata.

| Characteristic | All Patients (n = 342) |
| --- | --- |
| Age (years), median [IQR] | 54 (40 – 65) |
| Gender (Female/Male), n% | 48.39%, 51.61% |
| Magnetic Field Strength | 1.5 T (Siemens) |
| Clinical Standard Protocol | Segmented full stack (9-15 SAx slices) |
| Reduced Slice Protocol | 1 LAx + 3, 4, or 5 SAx slices |

Abbreviations: LAx – Long Axis; SAx – Short Axis; IQR - Interquartile Range

### 2.2 Slice Selection Criteria

We evaluated the reduced slice method using N = 3, 4 or 5 short axis slices. To account for the longitudinal shortening of the left ventricle during systole, we retrospectively selected N equally spaced slices that remained intracavitary throughout the cardiac cycle from the full-stack of SAx slices. If a selected slice failed to contain a left ventricular endocardial segmentation at any cardiac phase, the selection was iteratively shifted inward to the next adjacent slice. If the adjustment resulted in less than 5 intracavitary slices, the patient was excluded for insufficient coverage.

### 2.3 Automatic definition of the atrioventricular plane

To define the anterior and inferior insertion points, we identified the basal-most points in the automatic long-axis segmentation. These points had the largest perpendicular distance to the mid-ventricular SAx slice, and the plane with the minimum angular deviation from the mid-ventricular slice passing through these two basal-most points was defined as the atrioventricular plane.

### 2.4 Volume Calculation

The current work builds upon the method as proposed in (9), which used a trapezoidal approximation for the shape of long-axial cross-section of the left ventricle of the heart. Here, we propose to use a prismoidal volume to approximate the volume between the most basal slice and the atrioventricular plane. The equations used for calculating the volumes are presented in the Appendix. In Fig. 2, points B, D, Q, R, S, P, C and A represent points along the LAx contour. 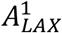 represents the LAx area enclosed between the points C, D, Q, and P, 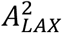 represents the LAx area enclosed between the points P, Q, R, and S, and 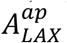 represents the LAx area below the apical-most SAx slice respectively. *A*_*Top*_ represents the area of top-most region which is closest to the atrioventricular junction, *A*_*Basal SAX*_ represents the area enclosed by the endocardium segmentation curve on the basal-most SAx slice, *A*_*Mid SAX*_ represents the area of segmentation curve on the middle SAx slice, and *A*_*Apical SAX*_ represents the area of the segmentation curve on the apical-most slice respectively. Similarly, *d*_*Top*_, *d*_*base*_, *d*_*mid*_ and *d*_*ap*_ represent the diameters of the almost circular regions of the segmented contours in the estimated atrio-ventricular plane, the basal-most, mid-ventricular and the apical-most SAx slices respectively. Here, *h* represents the distance between the basal-most SAx slice and the estimated top-most plane. *A*_*m*_is estimated using Eq. 4 in the Appendix.

**Figure 2:**
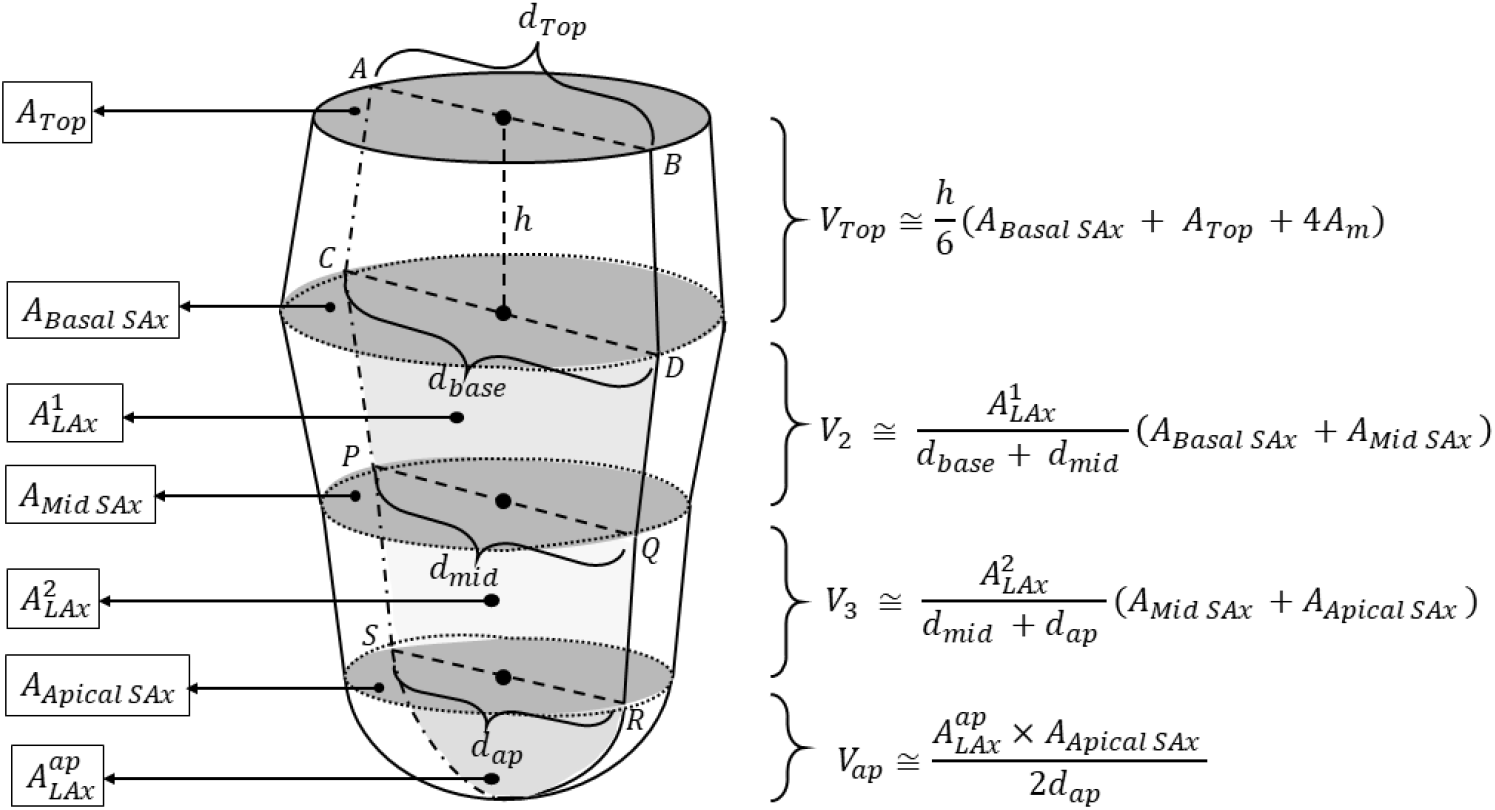
Schematic representation for volume calculation of different sections of the heart using 3 SAx slices and one LAx slice. The proposed model uses a prismoidal approximation for the volume between the basal-most slice and the atrioventricular plane.

### 2.5 Evaluation

For each patient, we evaluated all permutations of 3, 4, and 5 SAx slices with 2, 3, or 4-chamber LAx slices. These measurements were compared against the volumes derived from the full stack. The segmentations used for the reduced-slice method were the same as the corresponding slices in the full-stack reference.

### 2.6 Statistical Analysis

#### 2.6.1. Two-Way Repeated Measures ANOVA

Interaction effects were assessed using two-way repeated measures ANOVA (11) using the *Pingouin* python package (12). The subject-specific variance was accounted for across all 9 permutations (*3* SAx slice densities *×* 3 LAx views). Generalized eta squared 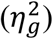, or effect size, is also reported along with the F-statistic and the Greenhouse-Geisser (13) corrected p-values.

#### 2.6.2. Post-hoc Comparisons

To test whether the change in measured volumes was statistically significant between *N* = 3 to *N* = 5 SAx slices, pairwise t-tests and Bayesian paired-samples t-test were performed. Bonferroni correction was used for multiple comparisons.

#### 2.6.3. Lin’s Concordance Correlation Coefficient and Intraclass Correlation Coefficient

Lin’s Concordance Correlation Coefficient (CCC) (14) and the Intraclass Correlation Coefficient (ICC) (15) with 95% confidence intervals for different parameters and different LAx and SAx combinations were used to assess the agreement and reliability between the full-stack volumes and the volumes calculated using the proposed method. Lin’s Concordance Correlation Coefficient was used as the primary measure of agreement between methods. Since same contours were used for both full-stack and the proposed reduced slice method, ICC (3,1) values representing a two-way mixed-effects model for a single measurement were reported. To establish the threshold for clinical certainty and facilitate comparisons with existing studies, the minimum detectable change (MDC) for different parameters is reported using the Bland-Altman LoAs, where MDC = (Upper LoA – Lower LoA)/2.

## 3 Results

Scan-time reductions for different long and short axis combinations averaged across all patients are shown in Table 2.

**Table 2:** Reduction in scan time across different long axis and short axis slice combinations. It is measured as a relative reduction between the total number of SAx and LAx slices acquired during the clinical examination and the number of slices used to carry out the analysis of the proposed method per patient. The reduction values reported here are averaged across different patients as the number of short-axis slices varies per patient.

| LAx and SAx combination | Average reduction in scan time (%) |
| --- | --- |
| 1 LAx + 3 SAx | 73 |
| 1 LAx + 4 SAx | 67 |
| 1 LAx + 5 SAx | 60 |

Abbreviations: LAx – Long Axis; SAx – Short Axis

As shown in Fig. 3, there is a systematic underestimation of EDV, ESV, and SV across all LAx views. In addition, the 3-chamber LAx view shows a larger negative bias between -11 to -26 milliliters as compared to the 2-chamber (bias between 0 and -12 mL) and 4-chamber views (bias between -6 and -17 mL). The plots in Fig. 3(d) for the EF show that the EF values are marginally overestimated (between 0 and 1.5 %) across all LAx and SAx combinations. The LoAs for EF were ± 7.5%.

**Figure 3:**
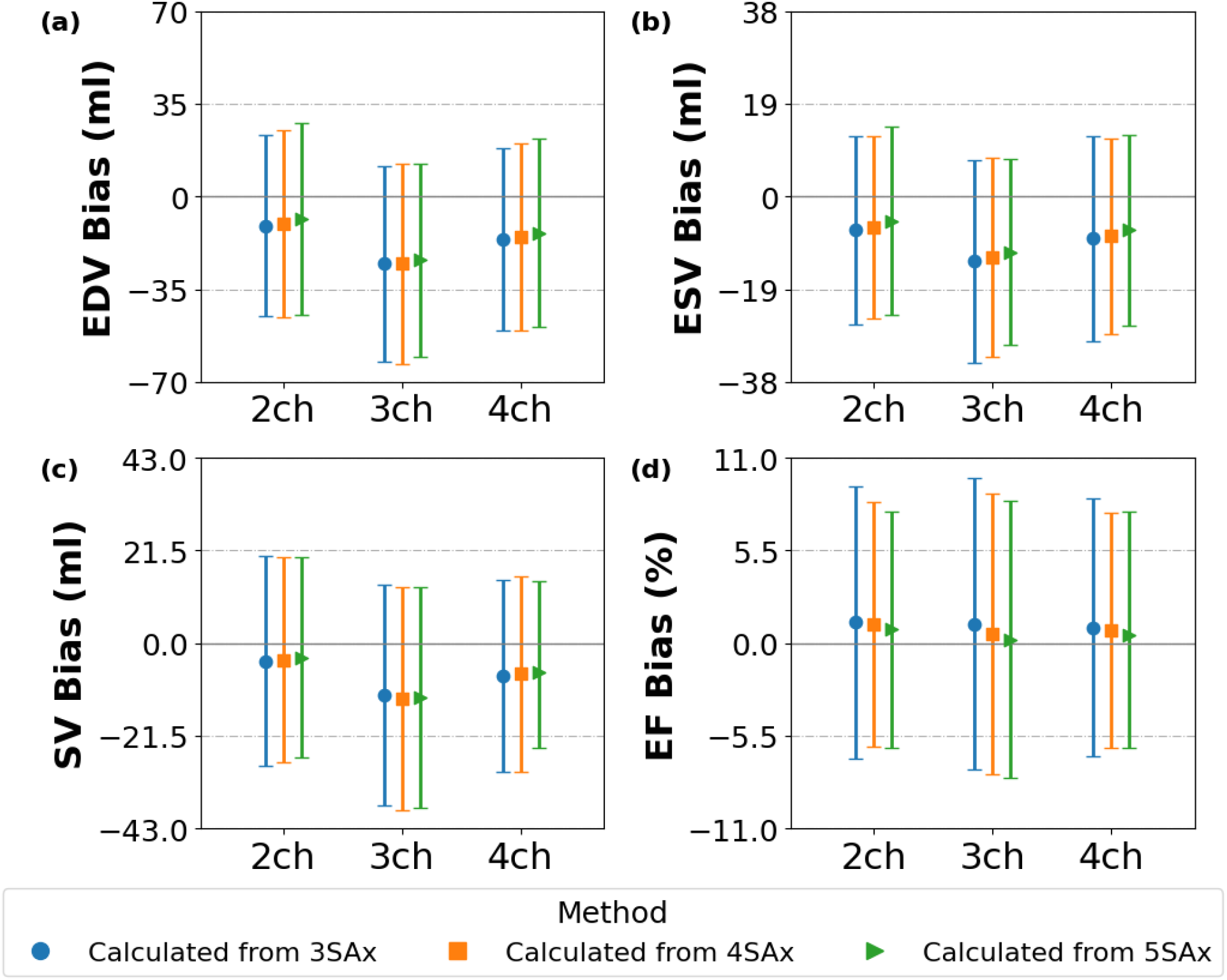
Bias and Limits of Agreement (LoA) plotted for different parameters (a) EDV, (b) ESV, (c) SV and (d) EF for different LAx and SAx slice combinations. The x-axis represents the different LAx views. For exact values, please refer to Table S1 in the Supplementary Materials section.

The CCC and ICC values for different parameters and different LAx and SAx combinations are shown in Fig. 4. These plots are used to assess the agreement and reliability between the full-stack volumes and the volumes calculated using the method proposed here. From the four subfigures, the CCC and ICC values for the 2ch + *N* SAx combinations are consistently the highest for EDV and ESV. The 2-chamber and 4-chamber combinations were comparable for SV while the 4ch + *N* SAx combinations gave higher agreements for EF. The 3-chamber LAx view appears to be the worst choice, with CCC for EDV and SV below 0.8, and a wider confidence interval, as shown in Fig. 4(a) and Fig. 4(c) respectively.

**Figure 4:**
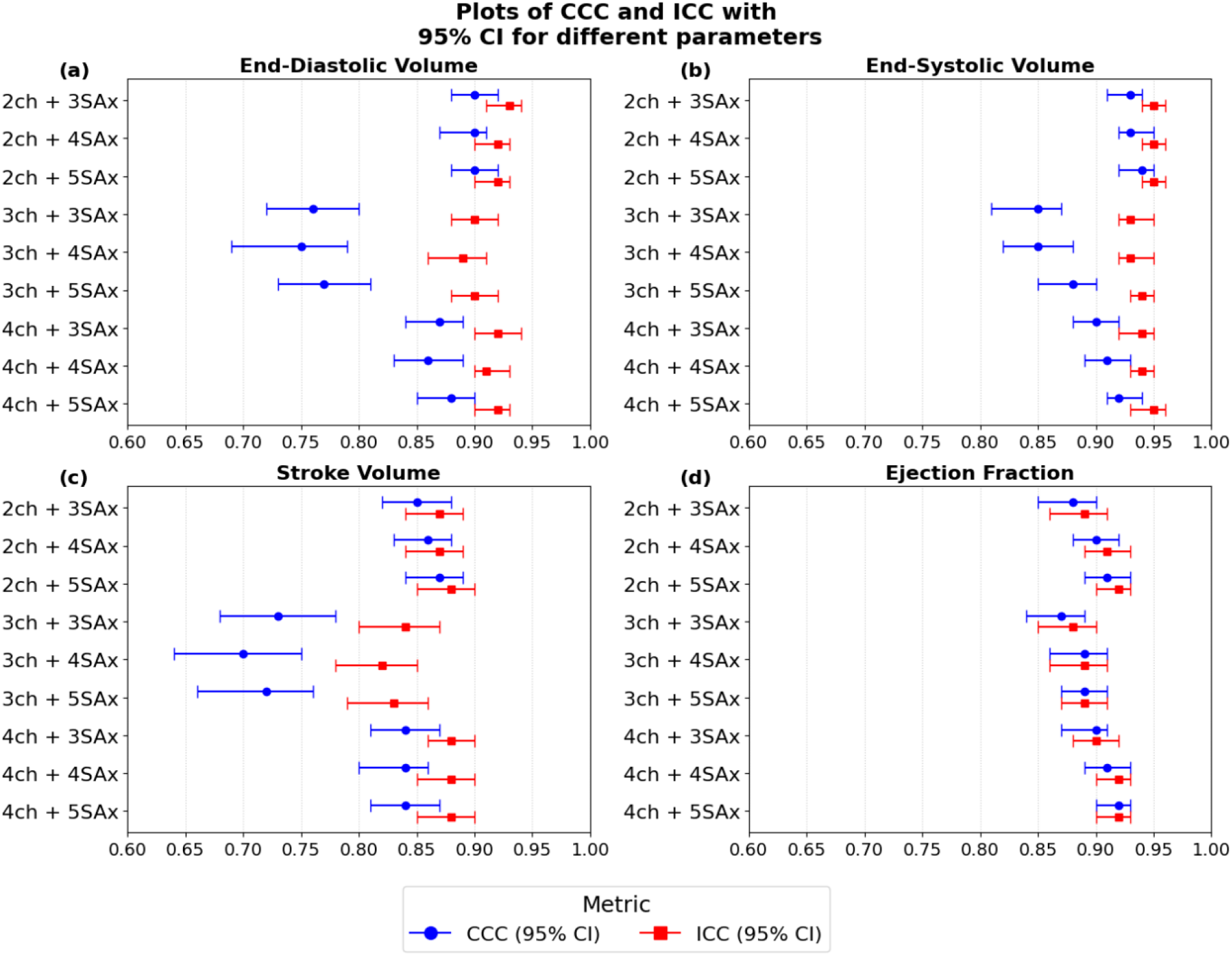
CCC and ICC graphs along with their confidence intervals for different parameters (a) EDV, (b) ESV, (c) SV and (d) EF plotted for different combinations of LAx and SAx slices. 2ch + 3SAx means a 2-chamber LAx along with 3 SAx slices are used for obtaining the volumetric parameters.

The results of the two-way measures ANOVA performed to assess the impact of two main factors, LAx view (2, 3, and 4-chamber) and the number of SAx slices (N = 3, 4, 5), on the volumetric parameters EDV, ESV, SV and EF are shown in Table S2. The choice of the LAx view was a significant driver for EDV, ESV, and EF. Meanwhile, the SAx slice count had a significant effect on EDV and ESV (both p<0.001). It is worth noting that the Effect Size 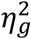 was always larger for LAx views as compared to the number of SAx slices for EDV, ESV, and SV. For instance, the LAx effect size for EDV 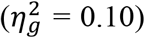 was significantly greater than the effect size of SAx slices for EDV 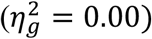 . These results indicate that the choice of LAx view was the more dominant factor in determining the accuracy of measurements. Conversely, EF displayed a different trend where the SAx slice count became a significant factor (p<0.001), whereas the LAx view was not (p = 0.079). Post-hoc pairwise comparisons revealed that for absolute volume measurements (EDV and ESV), a 5-SAx slice protocol was significantly better (p<0.001) than both 3 and 4 slice approaches across all LAx views, supported by decisive evidence from Bayesian analysis (2.66 ≤ log_10_(BF_10_) ≤ 10.57). In all these cases we did not find improvement by increasing the number of SAx slices from 3 to 4. For the functional parameters (SV and EF), accuracy did not improve with increasing number of SAx slices.

Extensive statistical results for all parameters and LAx and SAx combinations, including ANOVA effect sizes and pairwise post-hoc comparisons, are provided in the Supplementary Material (Tables S2 and S3). Bland-Altman plots for the agreement assessment between the proposed methodology and the full-stack method for different ventricular parameters is shown in Figures S1 – S9.

## 4 Discussion

In this study, we investigated a method for approximating the left ventricular volumes using a reduced number of slices, with only marginal loss of accuracy for SV and EF. Unlike (9), this study establishes a comparison between the reduced-slice method and the full-stack volume calculated automatically by the Segment software after the short-axis image stack segmentation. In addition, this study incorporates the volume between the basal-most slice used for volume calculation and the atrioventricular plane. A standard cine full-stack SAx acquisition acquires between 9 to 15 slices, which translates to 9 to 15 breath holds (often taking 3 to 6 minutes with rest in between) in addition to the three LAx slices. By reducing the protocol to 5 or fewer SAx and the LAx slice, we can cut the number of breath-holds significantly. This has the potential to minimize exhaustion among dyspneic or heart-failure patients while subsequently reducing the chances of motion artifacts or misalignment of SAx and LAx slices due to poor breath hold.

The results indicate a systematic underestimation of the LV volumetric parameters EDV, ESV, and SV when using the reduced-slice method for volume calculation as opposed to the full-stack method. We observe this bias primarily because of the geometric assumptions of the model. In addition to the geometric assumptions discussed above, this approach assumes the LAx contours to be uniformly distributed around a central axis which allows us to derive and use equations 1 to 3. Our results show that the volume calculations performed using 2-chamber LAx views yielded the most reliable results, probably because it better approximates a uniform contour around a central axis as compared to the other LAx views. Implementation of dynamic slice selection to account for the through-plane motion of the heart for basal-most and axial-most slice selection also makes the volume calculation sensitive to the position of these slices. A multicenter study by Bhuva et al. (6) reported the following scan-reproducibility results for EDV with a mean bias of 10 ml (LoA: -25 to 28ml) for automated analysis on full short axis cine stack. In comparison, the reduced-slice approach using 2ch + 5SAx slices had a comparable mean bias for EDV of -8 ml (LoA: -45 to 28ml). Similarly, the authors reported a mean bias of 10ml (LoA: -24 to 29ml) for automated analysis of SV and in comparison, our results for SV using the 2ch + 5 SAx protocol demonstrated a mean bias of -3 ml (LoA: -27 to 20ml). The EF for the proposed protocol was 0.83% (LoA: -6 to 8%) and an MDC of 7% as opposed to 4 % (LoA: -11 to 12%) with an MDC of 9% for the automated analysis, as reported in (6). With the LoAs for SV and EF being within the LoA for scan-rescan variability, this method can be used to reduce the number of breath holds without compromising accuracy in a clinical setting. Notably, the bias and LoA for SV and EF are below and within scan-rescan variability for 3 SAx with 2-chamber LAx, and by using 5 SAx this is also extended to the absolute volume metrics ESV and EDV, at the expense of two additional breath holds.

While the EDV, ESV, and SV showed a systematic underestimation bias, the EF values showed relatively low bias (0 – 1.5%) as shown in Fig. 3(d). This suggests that bias appears in the volume measurements throughout the cardiac cycle using the proposed method and gets mathematically neutralized by systematic bias cancellation when calculating the ratio of volumes, thereby maintaining the method’s clinical utility for assessing the global function. We also observe a trend of decreasing bias with an increase in the number of SAx slices. This trend, though visible, looks marginal in comparison to the decrease in bias when using 2 or 4-chamber LAx views instead of the 3-chamber LAx view.

The post-hoc pairwise comparisons revealed that for absolute volume measurements (EDV and ESV), a 5 SAx slice protocol was better than both 3 and 4 SAx approaches, regardless of the choice of LAx view. Contrary to this, for the functional parameters (SV and EF), we did not find an improvement in accuracy depending on the number of SAx slices, hinting that a reduced slice density is enough for the global LV function despite systematic underestimation of absolute volumes.

The results reveal that while addition of more SAx slices does statistically reduce the bias (p<0.001), the small effect size 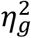 tells us that despite the improvement being statistically significant, it is clinically marginal compared to selection of the correct LAx view. Results in table S3 support this. The comparison between 3 and 5 SAx slices for 2-chamber (p<0.001, log_10_(BF_10_) = 6.42) and 3-chamber (p = 0.012, log_10_(BF_10_) = 1.01) view for the EDV shows that a change in LAx view had a greater influence on the results than the number of SAx slices.

The 3-chamber LAx demonstrated the highest negative bias and the lowest concordance among the long-axis orientations. A noticeable increase in LoAs for EDV with 3-chamber LAx makes it less suitable for LV quantification using this model. This is likely caused by the atrioventricular points being automatically selected based off the automatic myocardial segmentation in the LAx image, where a septal point is being used rather than the true annular position. This could potentially be mitigated by an alternative automatic segmentation approach to track the true annular point using a landmark based approach instead.

The generalized prismoidal formula was chosen to calculate the volume *V*_*Top*_ because it provides a higher-order geometric approximation of the basal annular region. This allows us to account for curvilinear nature of the basal myocardium as it approaches the mitral valve. The necessity of estimating *A*_*m*_ through diameter averaging is one of the contributors to the systematic bias visible in the plots of Fig. 3. The Standard Simpson’s rule (16) requires SAx slices to estimate the volume within the heart and in the absence of a physical slice above the basal-most slice, the generalized prismoidal formula was used to estimate the volume. Another source of error in this method comes from treating the intersection between the left ventricle and the atrioventricular plane as a circular region. Since only the two mitral annular insertion points *A* and *B* (Fig. 5(a)) are available per LAx view, fitting a circle is one of the feasible approaches. Literature shows that the mitral valve annulus is either an elliptical or D-shaped non-planar structure (17), (18) rather than circular, and this approximation may contribute to a systematic bias in the measurements, for EDV and ESV.

**Figure 5:**
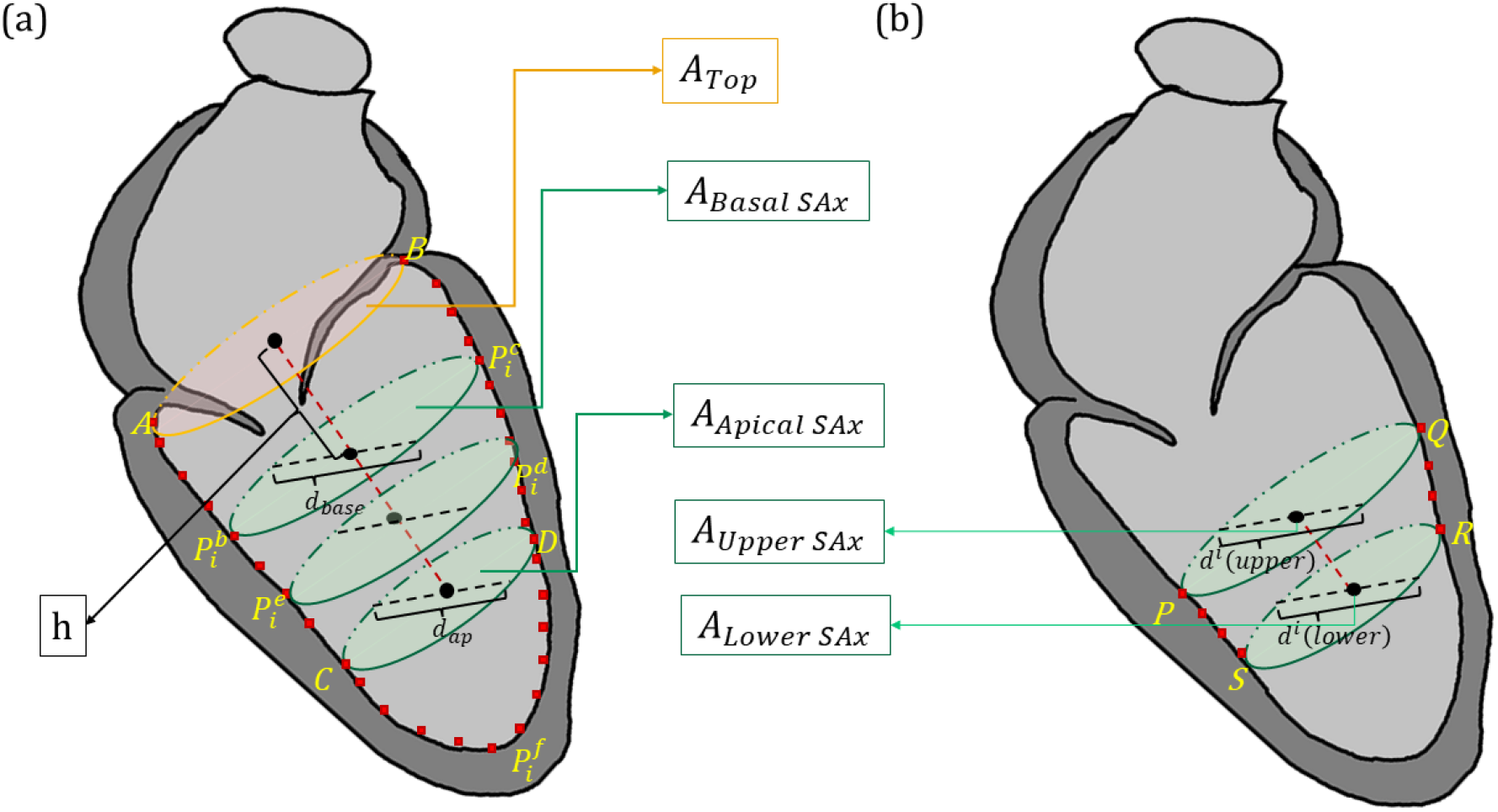
Two-chamber cross-sectional view of the heart. The red dots represent the points along the endocardial segmentation contour of the LAx view. Yellow text labels points on the segmentation contour. (a) Represents the regions above the basal-most slice and below the apical-most slice. The SAx slice in the middle represents the position of an arbitrary slice lying between the basal-most and apical-most slice. (b) Shows the points to be considered for volume calculation of the *i*^*th*^ heart section.

This method has the potential to be used as a rapid alternative where standard full-stack imaging is unfeasible, rather than a total replacement for the existing clinical CMR protocols. Furthermore, the reduced slice method can be used in conjunction with CMR methods that operate under time constraints to resolve arrhythmias in a clinical setting.

### 4.1 Limitations

The reduced-slice volumetric model offers reduced scan time but is limited by the systematic underestimation of absolute volumes. The patient cohort consists of unselected patients referred to a CMR and thus represents a mix of cardiovascular pathologies. Nevertheless, there is a potential risk that some diseases are not well represented by the material and thus might not work as well with the method based on heart geometries. In particular, the Karolinska University Hospital is a tertiary hospital that provides highly specialized health services, so the patient population may not accurately reflect the populations at primary and secondary health centers. This leaves a potential risk of selection bias for the generalizability of the results in this study. It is also important to mention that the exclusion of patient data where less than 5 SAx slices remained intracavitary throughout the cardiac cycle creates a selection bias towards larger hearts, or hearts with reduced longitudinal function. Patients with fewer than 5 SAx slices at end systole were excluded to maintain a uniformvalidation set. A prospective validation, where five equidistant slices can be prescribed regardless of heart size remains as a part of future work. A noticeable increase in LoAs for EDV with 3-chamber LAX makes it less suitable for LV quantification using this model. Implementation of dynamic slice selection to account for the through-plane motion of the heart for basal-most and axial-most slice selection makes the volume calculation sensitive to the position of these slices.

### 4.2 Conclusion

Using the proposed method, the number of breath holds and the scan time can be reduced, while at the same time maintaining acceptable accuracy with bias and LoA below scan-rescan variability for stroke volume and ejection fraction for a scan time reduction of 73%. The choice of LAx view has a stronger impact on the accuracy than the number of SAx slices, and 2 chamber LAx view performed best. However, by using 5 SAx slices also the absolute volume metrics ESV and EDV have lower bias and narrower LoA than previously reported scan-rescan variability.

## 5 Appendix

For a given number *N* of short-axis slices, the long-axis segmentation is divided into *N* + 1 sections. Volume of the *i*^*th*^ section, which is between the two slices *upper* and *lower*, is given by equation 1:

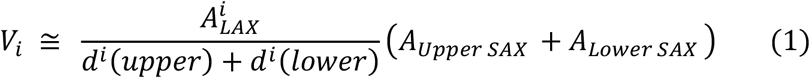

Where, *d*^*i*^*(upper)* and *d*^*i*^*(lower)* represent the diameter of the upper and lower short-axis slices encompassing the *i*^*th*^ section, *A*_*Upper SAX*_ and *A*_*Lower SAX*_ represent the areas bounded by the segmentation curve of the left-ventricle of the upper and lower short-axis slices respectively. 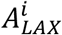 is the area encompassed by the points P, Q, R and S respectively, as shown in Fig. 5(b). The region 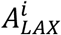 is approximated as a trapezoid. For the section below the apical-most slice, the volume is approximated using the equation 2 given below:

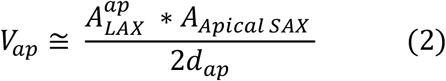

Where *A*_*Apical SAX*_ is the area of the segmentation curve present in the apical-most slice, 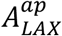 represents the area encompassed between the points C, D, and the long-axis segmentation curve and passing through the point 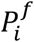 as shown in Fig. 5(a) lying below the apical-most slice, 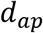 represents the diameter of the apical-most slice. The approach presented here makes use of the generalized prismoidal formula shown below:

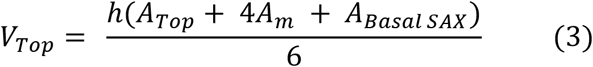

Here, *h* is the distance between the centroids of *A*_*Basal SAX*_ and the two topmost points *A* and *B, A*_*Basal SAX*_ is the area of the basal-most slice, *A*_*Top*_is the area calculated using the two topmost points as found in section 3.3 and shown in Fig. 5(a). Points *A* and *B* are treated as the end points of a diameter *d*_*Top*_ and *A*_*Top*_is calculated by approximating the annulus as a circular area. *A*_*m*_is the area of the region half-way between the atrio-ventricular plane and the basal-most slice. Since *A*_*m*_ is unknown, it is found by averaging the linear dimensions, not the areas. The middle diameter *d*_*m*_ is found using:

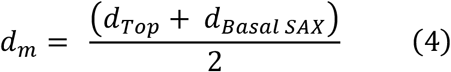

Additionally, the segmented endocardial regions are treated as circular with 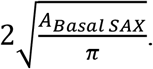 With this, 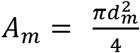 and one can estimate the atrio-ventricular volume *V*_*Top*_ using equation 3. Finally, the overall volume is estimated by summing up all the volumes calculated and is expressed by the following equation:

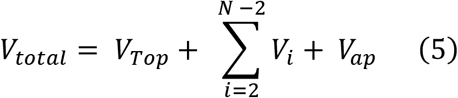

During slice planning in CMR, it is often not possible to get the LAx plane exactly perpendicular to the SAx plane. This is compensated for by multiplying the LAx area *A*_*Lax*_ with cos(*ϕ*) as shown in the following equation:

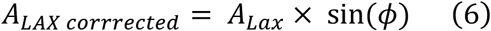

Where *ϕ* is the angle between the normal vectors of the SAx and LAx slices. Fig. 2 shows the application of the above-mentioned equations for three SAx slices and one LAx slice.

## Author Contributions

Aditya Tejaswi: Conceptualization, Methodology, Software, Validation, Formal Analysis, Data Curation, Writing - Original Draft, Writing – Review & Editing, Visualization, Alexander Fyrdahl: Conceptualization, Methodology, Resources, Writing – Review & Editing, Supervision, Project Administration, Funding Acquisition, Andreas Sigfridsson: Conceptualization, Methodology, Resources, Writing – Review & Editing, Supervision, Project Administration, Funding Acquisition

## Funding

Swedish Research Council (2022-04688, 2024-04452), Swedish Heart Lung foundation (20241479, 20251202), ALF Region Stockholm (RS2023-0859, RS2023 0872), Karolinska Institutet (2023-01434).

## Code Availability

The code used in this study is publicly available at Github (https://github.com/KarolinskaCMR/Reduced_Slice_Volume_Calculation). The version corresponding to this manuscript is archived under release v1.0.0.

## Data Availability

This article contains results generated after the analysis of CMR images obtained from patients in a clinical setting. The patient MRI data is not publicly available due to patient privacy, consent and ethical restrictions governed by the permits of this project. De-identified processed data supporting the representative figure may be made available by the corresponding author upon reasonable request and subject to institutional and ethical approval.

## Abbreviations

CMR: Cardiovascular Magnetic Resonance
LV: Left Ventricular
EF: Ejection Fraction
SAx: Short Axis
LAx: Long Axis
EDV: End Diastolic Volume
ESV: End Systolic Volume
SV: Stroke Volume
CCC: Concordance Correlation Coefficient
ICC: Intraclass Correlation Coefficient
ANOVA: Analysis of Variance
MDC: Minimum Detectable Change
LoA: Limits of Agreement
BF: Bayes Factor
AI: Artificial Intelligence
PACS: Patient Archiving and Communication System
SD: Standard Deviation

## Declarations

### Conflict of Interest

The authors have no competing interests to disclose.

### Ethical Approval

Data collection from the patients was carried out after getting approval from the Swedish Ethical Review Authority (DNR: 2011/1077-31/3 and 2022-01151-02), and all patients provided written informed consent.

## Notes

### Competing Interest Statement

The authors have declared no competing interest.

### Author Declarations

Data from the patient cohort for the current study was collected as part of the KAMRAT (Karolinska University Hospital's study of Magnetic Resonance Imaging of the Heart) studies. The study was approved by the Swedish Ethical Review Authority (DNR: 2011/1077-31/3 and 2022-01151-02) and all patients provided written informed consent.

### Summary of Updates

This revised version includes changes in response to the feedbacks received from reviewers. Minor textual and editorial changes have been made to improve readability and clarity. No new data or analyses have been added. The results and conclusion remain unchanged from the previous version.

