## Supplementary Material for "Left Ventricular Volume and Function Assessment Using a Reduced-Slice Approach in Cardiovascular Magnetic Resonance"

Table S1: Bias and Limits of Agreement for different parameters across different slice combinations.

| LAx Orientation | SAx slices | Parameter | Bias | LoA (lower) | LoA(upper) | Units |
|---|---|---|---|---|---|---|
| 2ch | 3 | EDV | -11.03 | -45.29 | 23.22 | ml |
| 2ch | 3 | ESV | -6.88 | -26.17 | 12.41 | ml |
| 2ch | 3 | SV | -4.16 | -28.47 | 20.16 | ml |
| 2ch | 3 | EF | 1.25 | -6.83 | 9.33 | % |
| 3ch | 3 | EDV | -25.45 | -62.23 | 11.34 | ml |
| 3ch | 3 | ESV | -13.32 | -34.08 | 7.44 | ml |
| 3ch | 3 | SV | -12.13 | -37.69 | 13.43 | ml |
| 3ch | 3 | EF | 1.14 | -7.51 | 9.8 | % |
| 4ch | 3 | EDV | -16.11 | -50.6 | 18.38 | ml |
| 4ch | 3 | ESV | -8.56 | -29.55 | 12.43 | ml |
| 4ch | 3 | SV | -7.55 | -29.75 | 14.65 | ml |
| 4ch | 3 | EF | 0.93 | -6.73 | 8.58 | % |
| 2ch | 4 | EDV | -10.17 | -45.47 | 25.13 | ml |
| 2ch | 4 | ESV | -6.31 | -24.92 | 12.31 | ml |
| 2ch | 4 | SV | -3.86 | -27.57 | 19.84 | ml |
| 2ch | 4 | EF | 1.13 | -6.1 | 8.37 | % |
| 3ch | 4 | EDV | -25.29 | -63.13 | 12.54 | ml |
| 3ch | 4 | ESV | -12.26 | -32.85 | 7.93 | ml |
| 3ch | 4 | SV | -12.84 | -38.68 | 13.01 | ml |
| 3ch | 4 | EF | 0.54 | -7.78 | 8.86 | % |
| 4ch | 4 | EDV | -15.24 | -50.51 | 20.03 | ml |
| 4ch | 4 | ESV | -8.1 | -28.15 | 11.95 | ml |
| 4ch | 4 | SV | -7.14 | -29.74 | 15.46 | ml |
| 4ch | 4 | EF | 0.78 | -6.18 | 7.75 | % |
| 2ch | 5 | EDV | -8.35 | -44.65 | 27.95 | ml |
| 2ch | 5 | ESV | -5.01 | -24.29 | 14.26 | ml |
| 2ch | 5 | SV | -3.34 | -26.61 | 19.94 | ml |
| 2ch | 5 | EF | 0.83 | -6.18 | 7.84 | % |
| 3ch | 5 | EDV | -23.91 | -60.37 | 12.55 | ml |
| 3ch | 5 | ESV | -11.38 | -30.44 | 7.69 | ml |
| 3ch | 5 | SV | -12.53 | -38.09 | 13.03 | ml |
| 3ch | 5 | EF | 0.23 | -7.99 | 8.45 | % |
| 4ch | 5 | EDV | -13.72 | -49.09 | 21.65 | ml |
| 4ch | 5 | ESV | -6.88 | -26.36 | 12.59 | ml |
| 4ch | 5 | SV | -6.83 | -24.29 | 14.26 | ml |
| 4ch | 5 | EF | 0.46 | -6.18 | 7.84 | % |

Table S2: Results for two-way repeated measures ANOVA. The table compares the effect of LAx view, number of SAx slices and the effect of both put together for the volumetric parameters. The reported p-values reported are Greenhouse-Geisser corrected and effect size represents generalized eta squared ($\eta_g^2$). df = degrees of freedom, GG-corr = Greenhouse-Geisser corrected

| Parameter | Source of Variation | F-statistic | df | p-value (GG-corr) | Effect Size |
|---|---|---|---|---|---|
| **EDV** | LAx View | 96.21 | 2 | < 0.001 | 0.1041 |
| **EDV** | Slice Count | 19.10 | 2 | < 0.001 | 0.0026 |
| **EDV** | LAx View * Slice Count | 5.99 | 4 | 0.010 | 0.0001 |
| **ESV** | LAx View | 58.89 | 2 | < 0.001 | 0.0646 |
| **ESV** | Slice Count | 47.66 | 2 | < 0.001 | 0.0059 |
| **ESV** | LAx View * Slice Count | 0.77 | 4 | 0.405 | 0.0000 |
| **SV** | LAx View | 89.38 | 2 | < 0.001 | 0.0789 |
| **SV** | Slice Count | 0.87 | 2 | 0.399 | 0.0002 |
| **SV** | LAx View * Slice Count | 8.32 | 4 | 0.001 | 0.0004 |
| **EF** | LAx View | 2.58 | 2 | 0.079 | 0.0023 |
| **EF** | Slice Count | 15.69 | 2 | < 0.001 | 0.0041 |
| **EF** | LAx View * Slice Count | 6.29 | 4 | 0.003 | 0.0007 |

Table S3: Results for post-hoc pairwise comparisons to evaluate the effect of different combinations of LAx views and SAx slices for different volumetric parameters. The p-values reported here are Bonferroni-corrected.

| Parameter | LAx Orientation | Comparison | p-value (Bonferroni-corr) | Bayes Factor $\log_{10}(BF_{10})$ |
|---|---|---|---|---|
| **EDV** | 2ch | 3 vs 4 SAx slices | 0.199 | -0.09 |
| **EDV** | 2ch | 3 vs 5 SAx slices | < 0.001 | 6.42 |
| **EDV** | 2ch | 4 vs 5 SAx slices | < 0.001 | 5.12 |
| **EDV** | 3ch | 3 vs 4 SAx slices | 1.000 | -1.18 |
| **EDV** | 3ch | 3 vs 5 SAx slices | 0.012 | 1.01 |
| **EDV** | 3ch | 4 vs 5 SAx slices | < 0.001 | 2.66 |
| **EDV** | 4ch | 3 vs 4 SAx slices | 0.189 | -0.07 |
| **EDV** | 4ch | 3 vs 5 SAx slices | < 0.001 | 6.82 |
| **EDV** | 4ch | 4 vs 5 SAx slices | < 0.001 | 4.97 |
| **ESV** | 2ch | 3 vs 4 SAx slices | 0.070 | 0.31 |
| **ESV** | 2ch | 3 vs 5 SAx slices | < 0.001 | 7.99 |
| **ESV** | 2ch | 4 vs 5 SAx slices | < 0.001 | 7.90 |
| **ESV** | 3ch | 3 vs 4 SAx slices | 0.001 | 1.87 |
| **ESV** | 3ch | 3 vs 5 SAx slices | < 0.001 | 10.57 |
| **ESV** | 3ch | 4 vs 5 SAx slices | < 0.001 | 6.10 |
| **ESV** | 4ch | 3 vs 4 SAx slices | 0.327 | -0.27 |
| **ESV** | 4ch | 3 vs 5 SAx slices | < 0.001 | 9.68 |
| **ESV** | 4ch | 4 vs 5 SAx slices | < 0.001 | 6.92 |
| **SV** | 2ch | 3 vs 4 SAx slices | 1.000 | -1.00 |
| **SV** | 2ch | 3 vs 5 SAx slices | 0.224 | -0.13 |
| **SV** | 2ch | 4 vs 5 SAx slices | 0.565 | -0.47 |
| **SV** | 3ch | 3 vs 4 SAx slices | 0.859 | -0.62 |
| **SV** | 3ch | 3 vs 5 SAx slices | 1.000 | -1.03 |
| **SV** | 3ch | 4 vs 5 SAx slices | 1.000 | -0.95 |
| **SV** | 4ch | 3 vs 4 SAx slices | 1.000 | -0.96 |
| **SV** | 4ch | 3 vs 5 SAx slices | 0.662 | -0.53 |
| **SV** | 4ch | 4 vs 5 SAx slices | 1.000 | -0.89 |
| **EF** | 2ch | 3 vs 4 SAx slices | 1.000 | -0.96 |
| **EF** | 2ch | 3 vs 5 SAx slices | 0.013 | 0.96 |
| **EF** | 2ch | 4 vs 5 SAx slices | 0.009 | 1.11 |
| **EF** | 3ch | 3 vs 4 SAx slices | < 0.001 | 2.41 |
| **EF** | 3ch | 3 vs 5 SAx slices | < 0.001 | 5.43 |
| **EF** | 3ch | 4 vs 5 SAx slices | 0.014 | 0.94 |
| **EF** | 4ch | 3 vs 4 SAx slices | 1.000 | -0.93 |
| **EF** | 4ch | 3 vs 5 SAx slices | 0.016 | 0.90 |
| **EF** | 4ch | 4 vs 5 SAx slices | 0.025 | 0.71 |

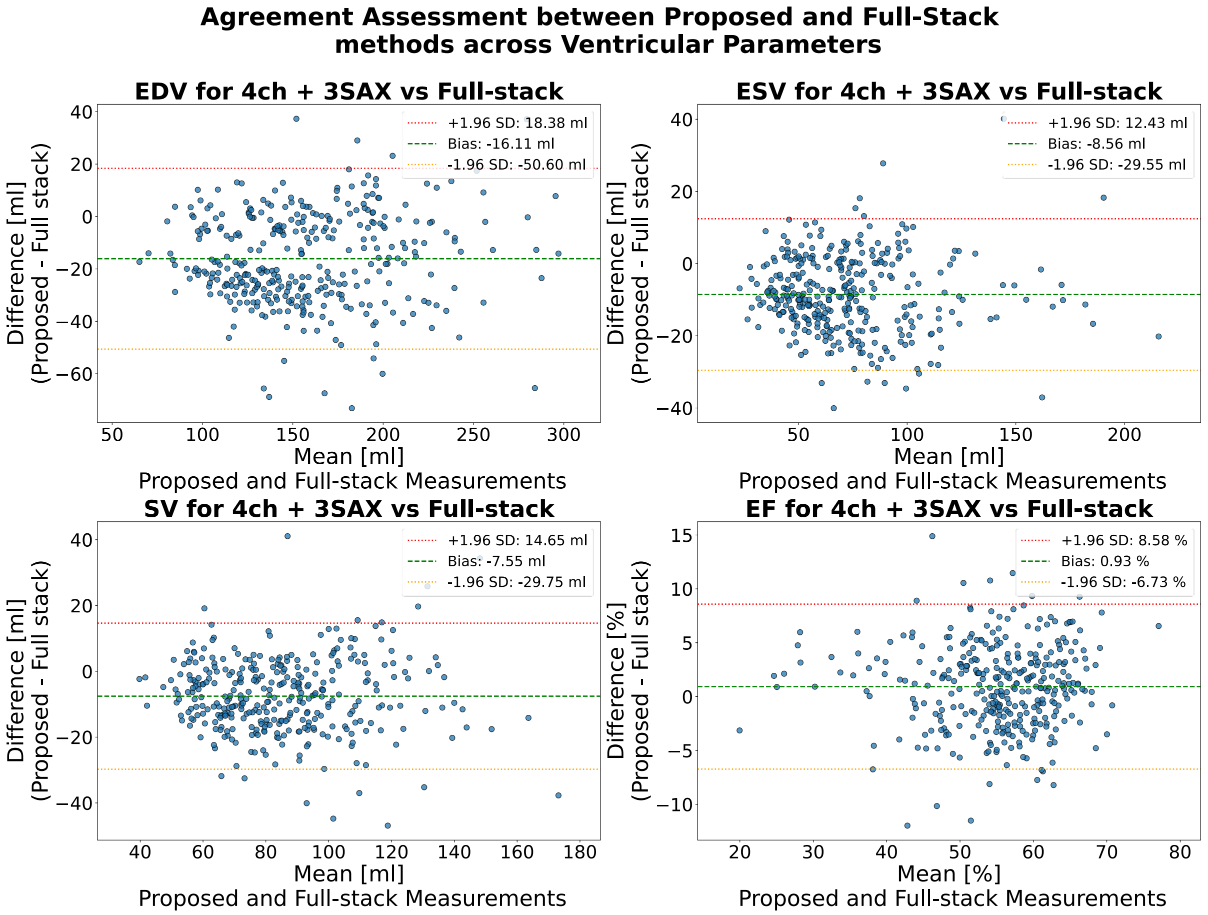


Figure S1: Bland-Altman agreement assessment for 4-chamber LAx view with 3 SAx slices. Scatter plots show the agreement between the proposed geometric method and the full-stack reference for End-Diastolic Volume (EDV), End-Systolic Volume (ESV), Stroke Volume (SV), and Ejection Fraction (EF). The x-axis represents the mean of the two methods, and the y-axis represents the difference (Proposed - Full Stack). The dashed green line indicates the mean bias, while the dotted red and orange lines represent the 95% limits of agreement (±1.96 SD).

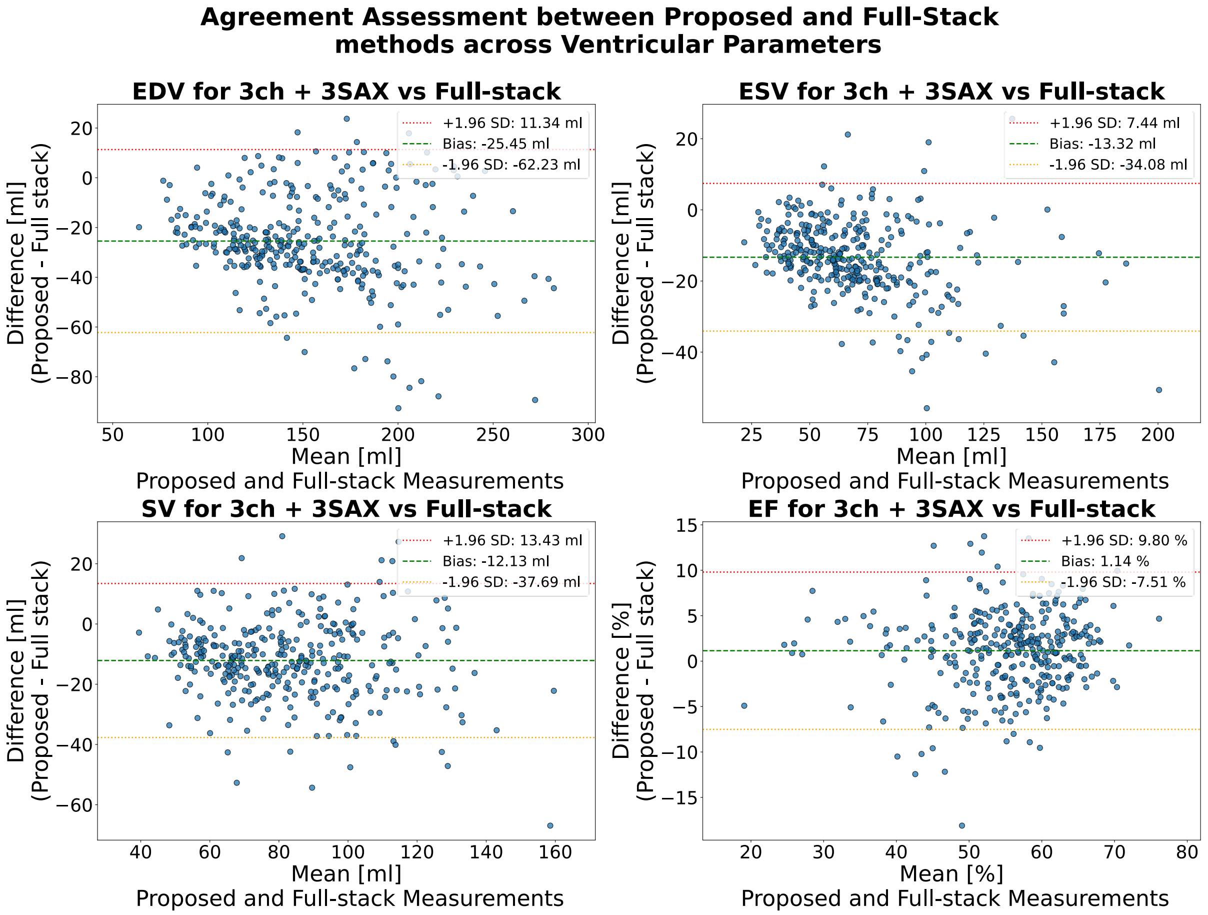


Figure S2: Bland-Altman agreement assessment for 3-chamber LAx view with 3 SAx slices. Scatter plots show the agreement between the proposed geometric method and the full-stack reference for End-Diastolic Volume (EDV), End-Systolic Volume (ESV), Stroke Volume (SV), and Ejection Fraction (EF). The x-axis represents the mean of the two methods, and the y-axis represents the difference (Proposed – Full Stack). The dashed green line indicates the mean bias, while the dotted red and orange lines represent the 95% limits of agreement (±1.96 SD).

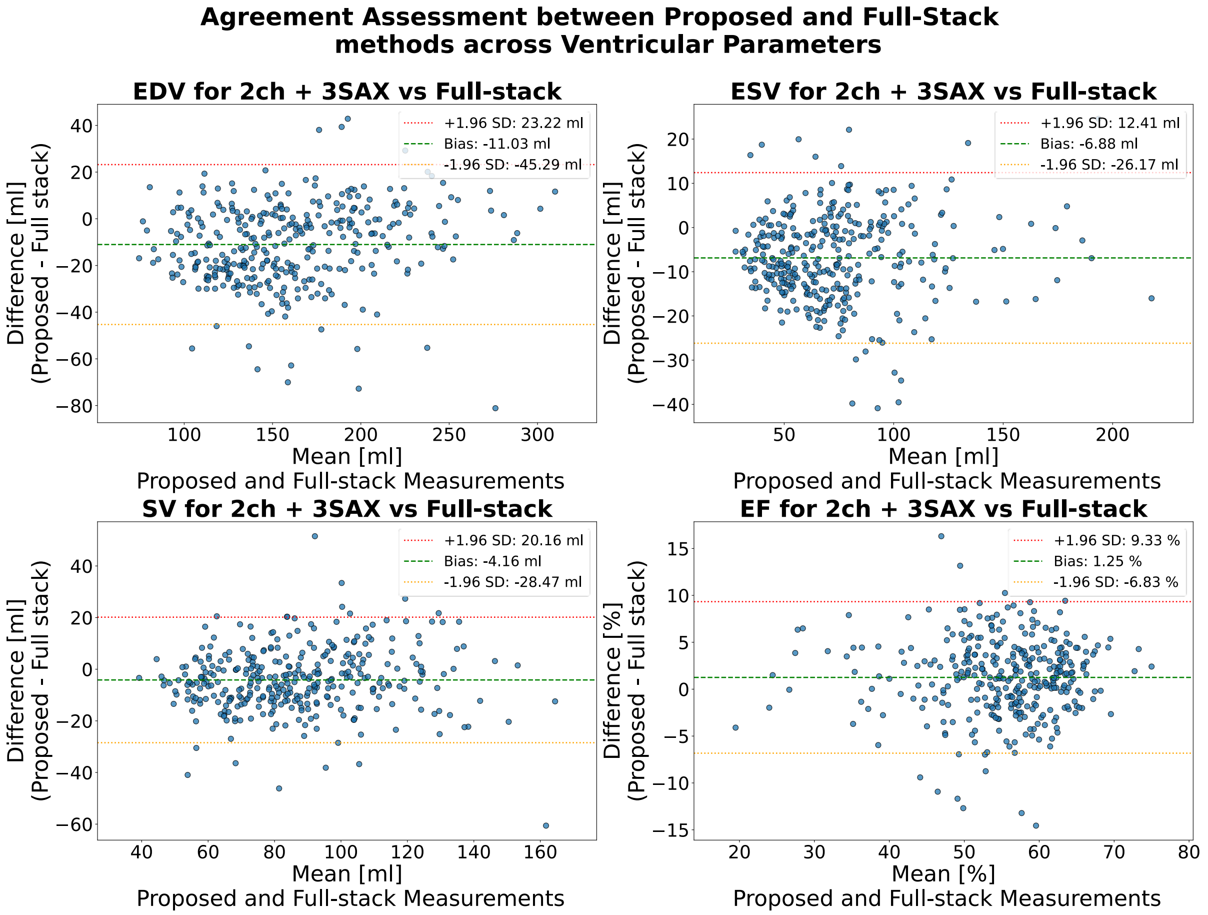


Figure S3: Bland-Altman agreement assessment for 2-chamber LAx view with 3 SAx slices. Scatter plots show the agreement between the proposed geometric method and the full-stack reference for End-Diastolic Volume (EDV), End-Systolic Volume (ESV), Stroke Volume (SV), and Ejection Fraction (EF). The x-axis represents the mean of the two methods, and the y-axis represents the difference (Proposed – Full Stack). The dashed green line indicates the mean bias, while the dotted red and orange lines represent the 95% limits of agreement (±1.96 SD).

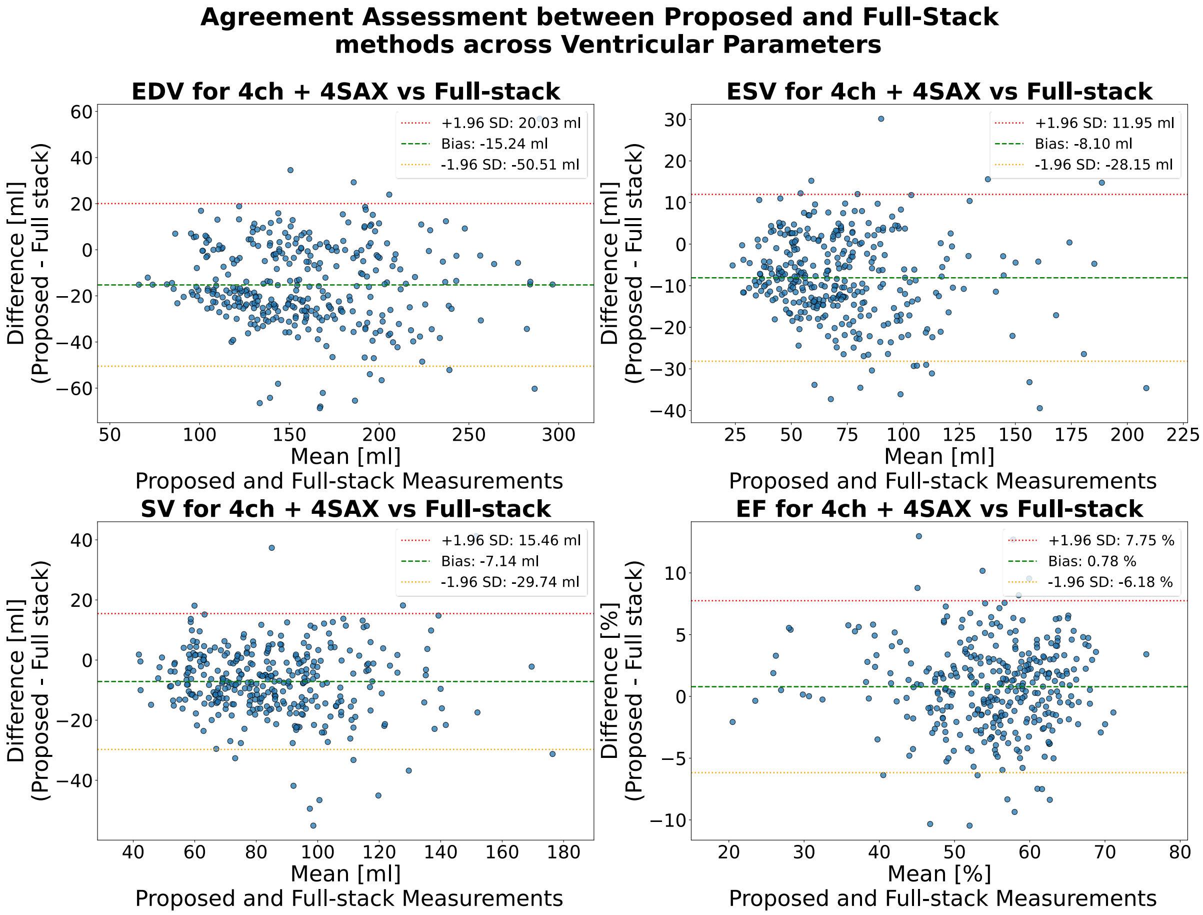


Figure S4: Bland-Altman agreement assessment for 4-chamber LAx view with 4 SAx slices. Scatter plots show the agreement between the proposed geometric method and the full-stack reference for End-Diastolic Volume (EDV), End-Systolic Volume (ESV), Stroke Volume (SV), and Ejection Fraction (EF). The x-axis represents the mean of the two methods, and the y-axis represents the difference (Proposed – Full Stack). The dashed green line indicates the mean bias, while the dotted red and orange lines represent the 95% limits of agreement (±1.96 SD).

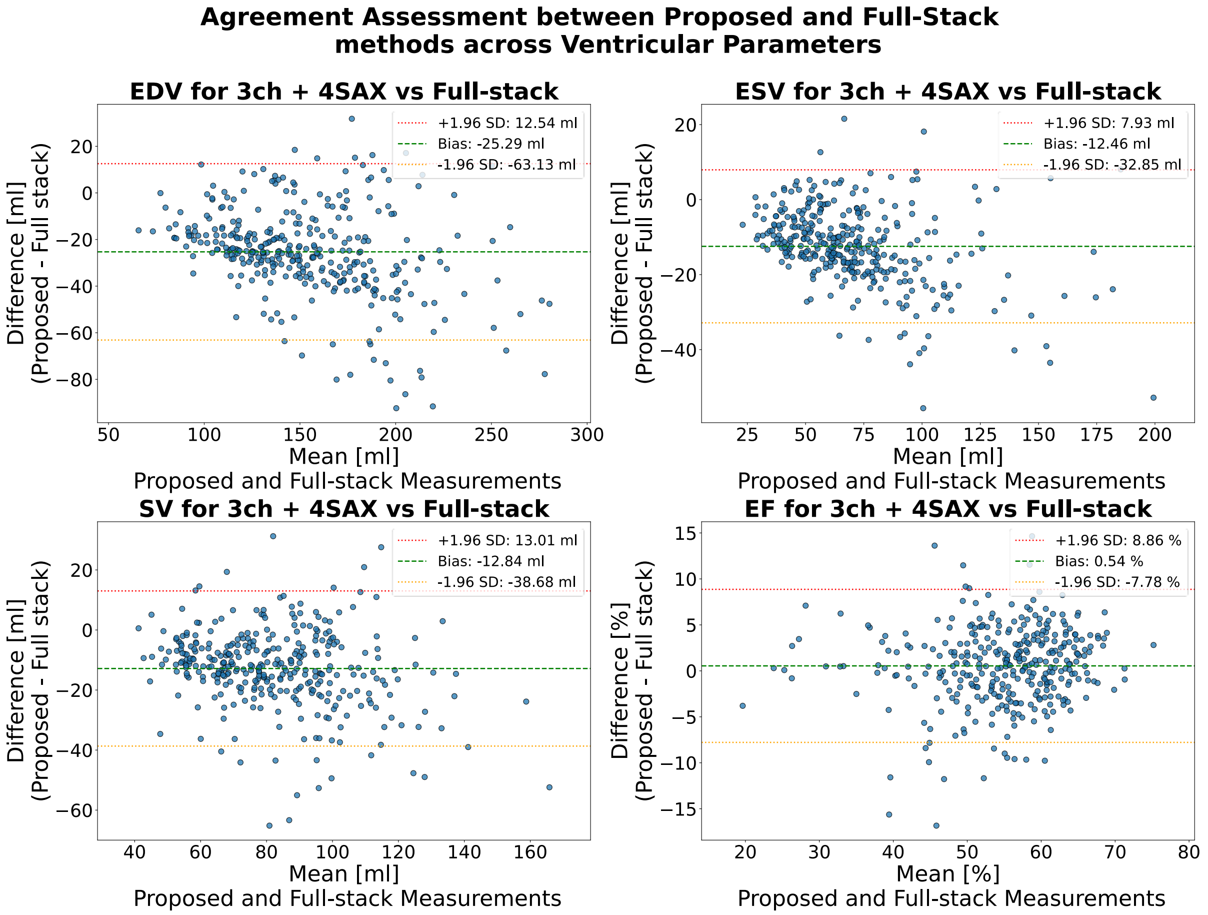


Figure S5: Bland-Altman agreement assessment for 3-chamber LAx view with 4 SAx slices. Scatter plots show the agreement between the proposed geometric method and the full-stack reference for End-Diastolic Volume (EDV), End-Systolic Volume (ESV), Stroke Volume (SV), and Ejection Fraction (EF). The x-axis represents the mean of the two methods, and the y-axis represents the difference (Proposed – Full Stack). The dashed green line indicates the mean bias, while the dotted red and orange lines represent the 95% limits of agreement (±1.96 SD).

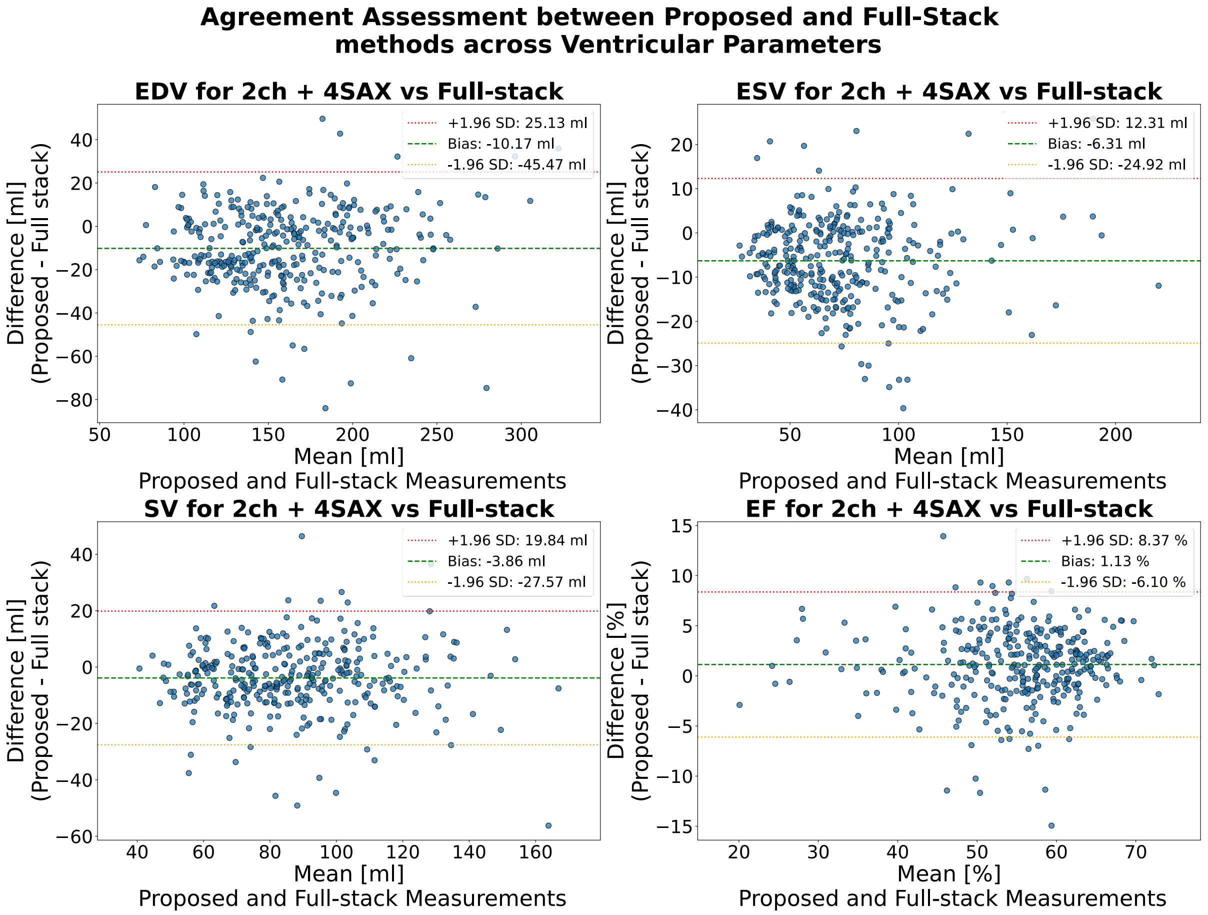


Figure S6: Bland-Altman agreement assessment for 2-chamber LAx view with 4 SAx slices. Scatter plots show the agreement between the proposed geometric method and the full-stack reference for End-Diastolic Volume (EDV), End-Systolic Volume (ESV), Stroke Volume (SV), and Ejection Fraction (EF). The x-axis represents the mean of the two methods, and the y-axis represents the difference (Proposed – Full Stack). The dashed green line indicates the mean bias, while the dotted red and orange lines represent the 95% limits of agreement (±1.96 SD).

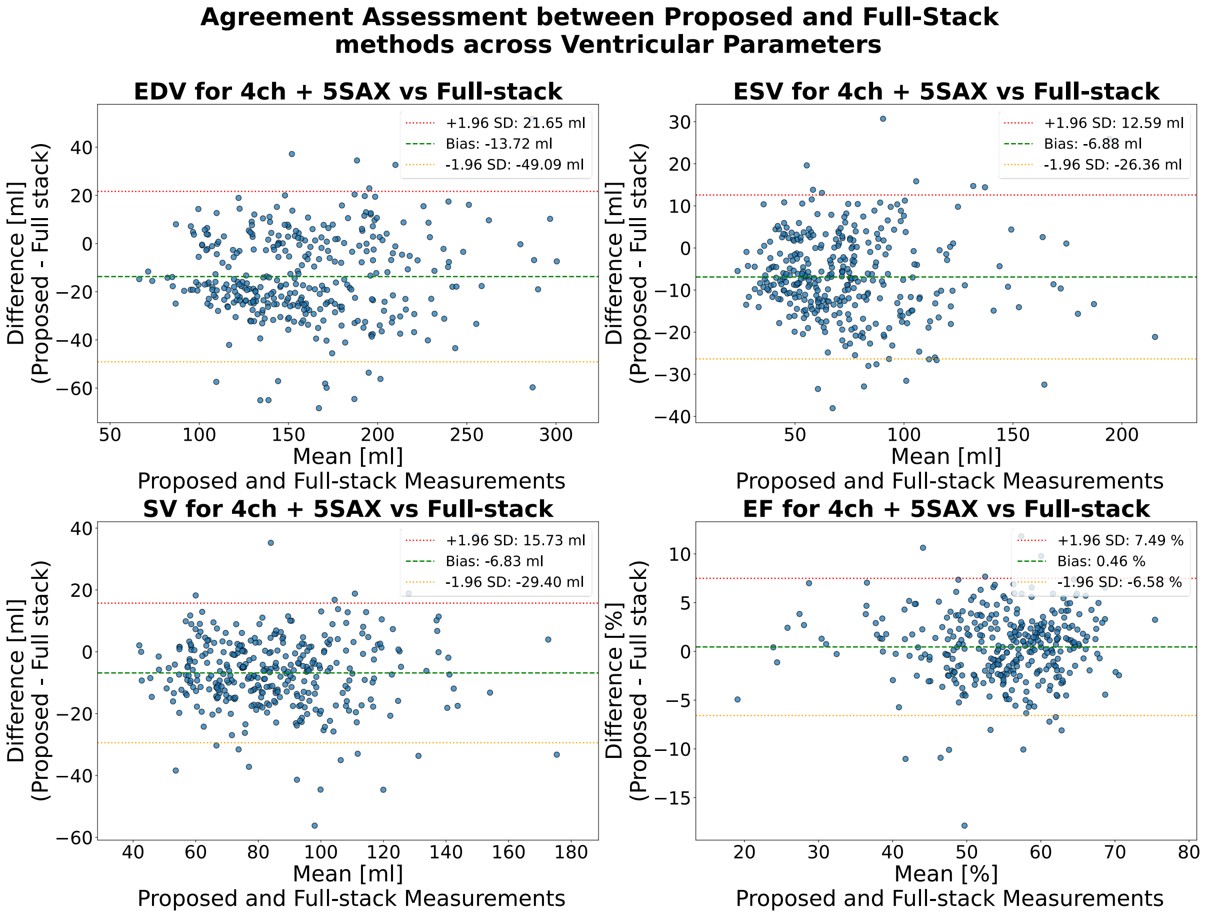


Figure S7: Bland-Altman agreement assessment for 4-chamber LAx view with 5 SAx slices. Scatter plots show the agreement between the proposed geometric method and the full-stack reference for End-Diastolic Volume (EDV), End-Systolic Volume (ESV), Stroke Volume (SV), and Ejection Fraction (EF). The x-axis represents the mean of the two methods, and the y-axis represents the difference (Proposed – Full Stack). The dashed green line indicates the mean bias, while the dotted red and orange lines represent the 95% limits of agreement (±1.96 SD).

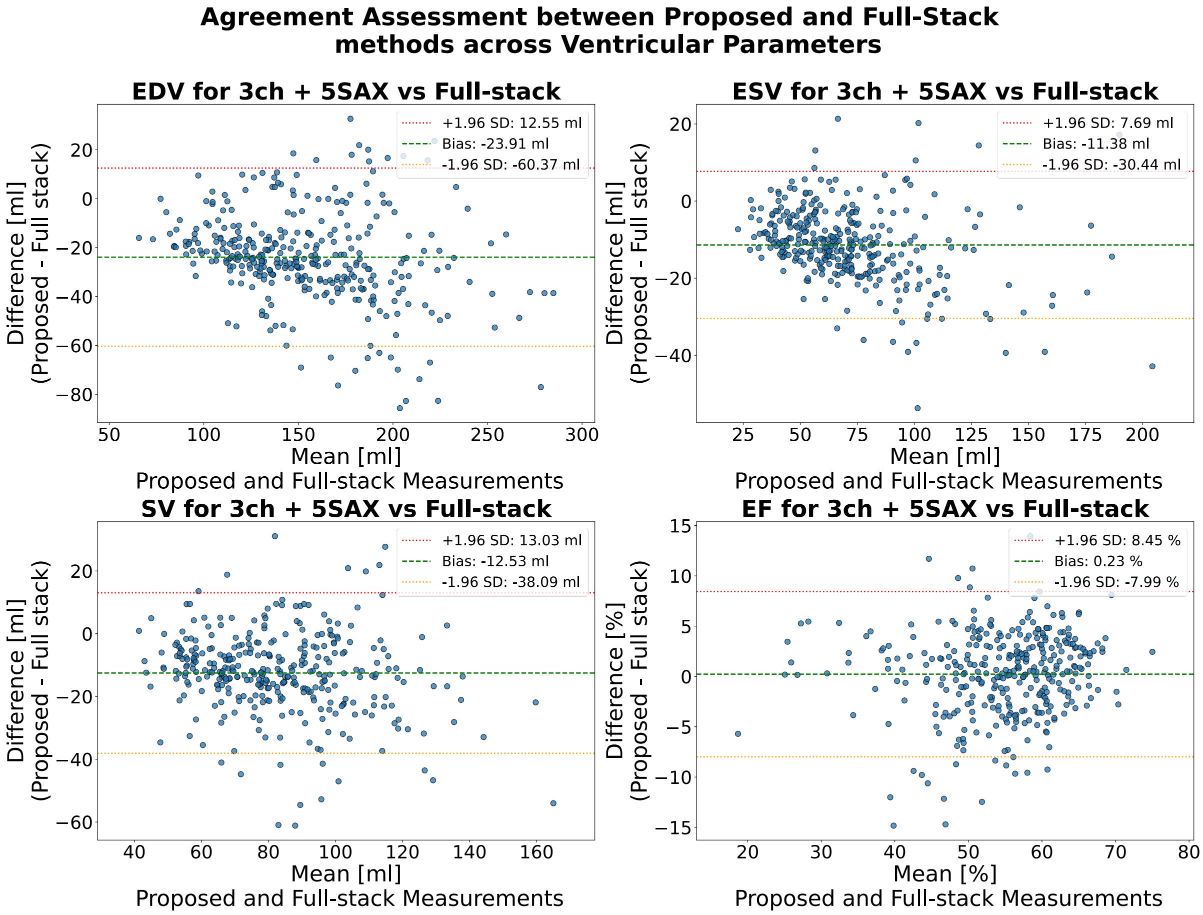


Figure S8: Bland-Altman agreement assessment for 3-chamber LAx view with 5 SAx slices. Scatter plots show the agreement between the proposed geometric method and the full-stack reference for End-Diastolic Volume (EDV), End-Systolic Volume (ESV), Stroke Volume (SV), and Ejection Fraction (EF). The x-axis represents the mean of the two methods, and the y-axis represents the difference (Proposed – Full Stack). The dashed green line indicates the mean bias, while the dotted red and orange lines represent the 95% limits of agreement (±1.96 SD).

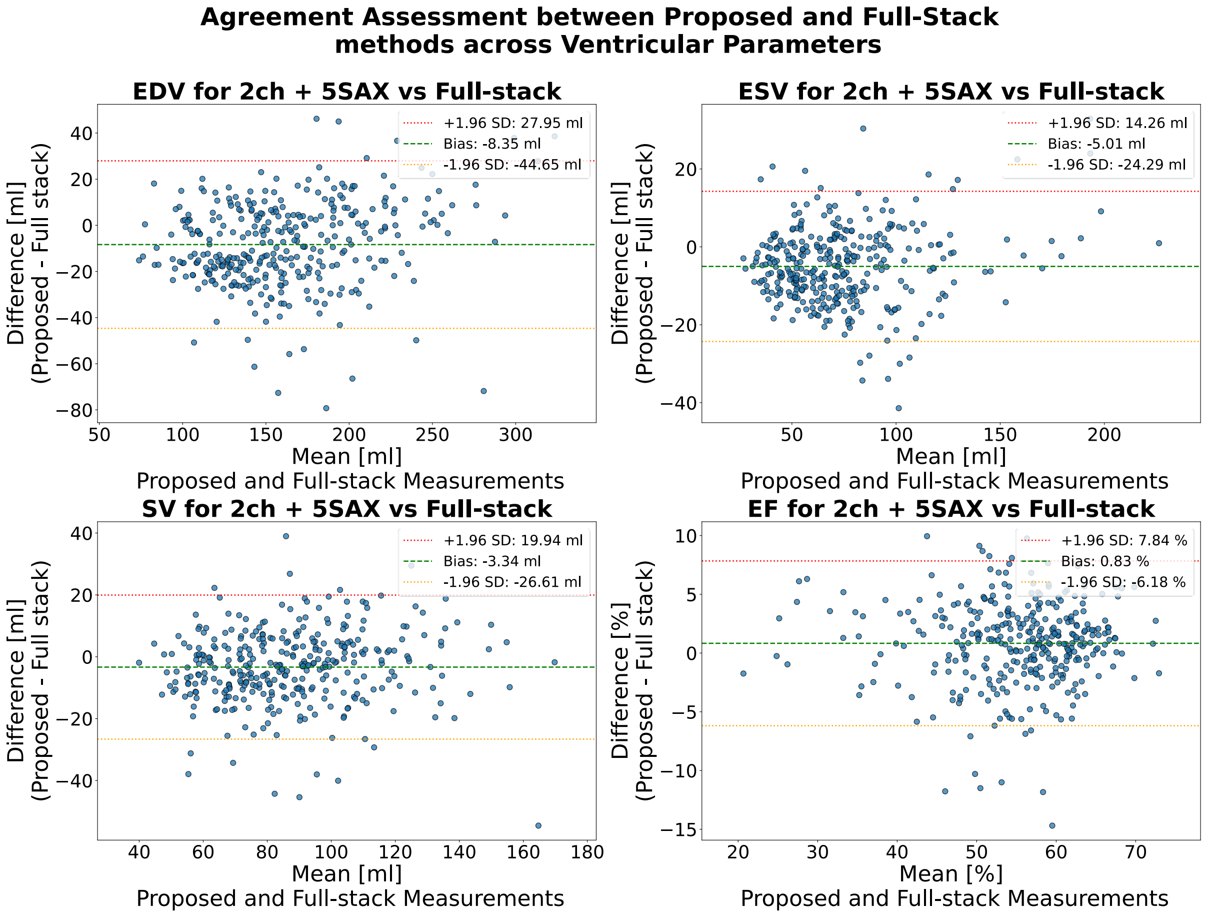


Figure S9: Bland-Altman agreement assessment for 2-chamber LAx view with 5 SAx slices. Scatter plots show the agreement between the proposed geometric method and the full-stack reference for End-Diastolic Volume (EDV), End-Systolic Volume (ESV), Stroke Volume (SV), and Ejection Fraction (EF). The x-axis represents the mean of the two methods, and the y-axis represents the difference (Proposed – Full Stack). The dashed green line indicates the mean bias, while the dotted red and orange lines represent the 95% limits of agreement (±1.96 SD).
